# A Pan-Cancer Analysis of Lesion-Level Treatment Response to Extend the ‘Seed and Soil’ Paradigm

**DOI:** 10.1101/2025.09.16.25335893

**Authors:** Dilley Ian, Zhou Jiawei, Li Quefeng, Cao Yanguang

**Affiliations:** Division of Pharmacotherapy and Experimental Therapeutics, UNC Eshelman School of Pharmacy, University of North Carolina at Chapel Hill, Chapel Hill, NC, 27599, USA; Lineberger Comprehensive Cancer Center, School of Medicine, University of North Carolina at Chapel Hill, Chapel Hill, NC 27599, USA; Department of Biostatistics, Gillings School of Global Public Health, University of North Carolina at Chapel Hill, Chapel Hill, NC, 27599, USA

## Abstract

**Background:** The classical “seed and soil” hypothesis suggests that metastatic spread is shaped by tumor-intrinsic traits (“seeds”) and the organ-specific microenvironment (“soil”). We expand this concept to explain lesion-level therapeutic responses and phenotypic variability across metastatic cancers.

**Methods:** We analyzed 55,220 lesions from 6,087 patients enrolled in 20 clinical trials across six cancer types. Using nonlinear mixed-effects modeling, we estimated lesion-specific parameters: regression rate (kkill), progression rate (kge), and resistant fraction (Fx). Multivariable Cox models, adjusted for cancer type, treatment modality, and clinical covariates, were used to assess organ-specific response patterns. Two key microenvironmental features—Vascular Perfusion and Leakiness Index (VaPLI) and tissue immune tolerance—were evaluated as predictors of lesion-level phenotypes.

**Results:** Treatment response dynamics varied significantly across metastatic sites, even within the same cancer type and therapy. Lesion-level responses reflected a strong seed–soil interaction, influenced by treatment modality. Liver metastases showed high initial regression but rapid progression, while bone lesions, especially in prostate cancer, exhibited more stable responses. VaPLI and immune tolerance status were significant predictors of lesion behavior.

**Conclusions:** Our findings support an expanded seed–soil framework that incorporates physiological and therapeutic context, enabling site-aware treatment strategies and refined patient selection for metastatic cancer therapies.

## Background

Metastasis remains the leading cause of cancer-related mortality. Despite notable advances in systemic therapies, the prognosis for patients with metastatic disease remains poor. This clinical reality is driven not only by the intrinsic properties of tumor cells but also by the heterogeneous microenvironments of the tissues in which metastatic lesions reside, making it nearly impossible to achieve curability. The classic “seed and soil” hypothesis, first proposed by Stephen Paget in 1889, provides a foundational framework for understanding the non-random dissemination of metastases across anatomical sites(1). According to this model, tumor cells (the “seeds”) can only form metastatic colonies in permissive microenvironments (the “soil”) that support their survival, proliferation, and outgrowth. Critically, the microenvironment shapes the clonal composition, phenotypic plasticity, as well as the therapeutic responsiveness of metastatic lesions(2,3). The “soil” consists of a complex ecosystem of stromal and immune cells, extracellular matrix components, vascular architecture, and biochemical signals. These features dynamically interact with disseminated tumor cells, influencing not only metastatic colonization but also treatment sensitivity(4).

Given these observations, we propose an extension of the “seed and soil” paradigm (**Figure 1A)** that encompasses not only metastatic dissemination but also lesion-level therapeutic response. In this extended framework, the therapeutic effect is largely shaped by both the biological characteristics of tumor cells (the “seeds”) and the unique stromal, vascular, and immune features of the host organ (the “soil”). These features likely operate across cancer types and therapeutic contexts. For instance, colorectal cancer (CRC) liver metastases often demonstrate deeper yet shorter-lived responses to therapy compared to lung metastases(2,5). Similarly, patients with pancreatic ductal adenocarcinoma (PDAC) who develop isolated lung metastases experience significantly improved outcomes compared to those with liver involvement(6). These clinical patterns suggest that the prognostic and therapeutic significance of the metastatic site, the “soil”, and its interaction with the types of tumor cells, the “seeds”, warrants closer investigation in the context of treatment response.

**Figure 1.**
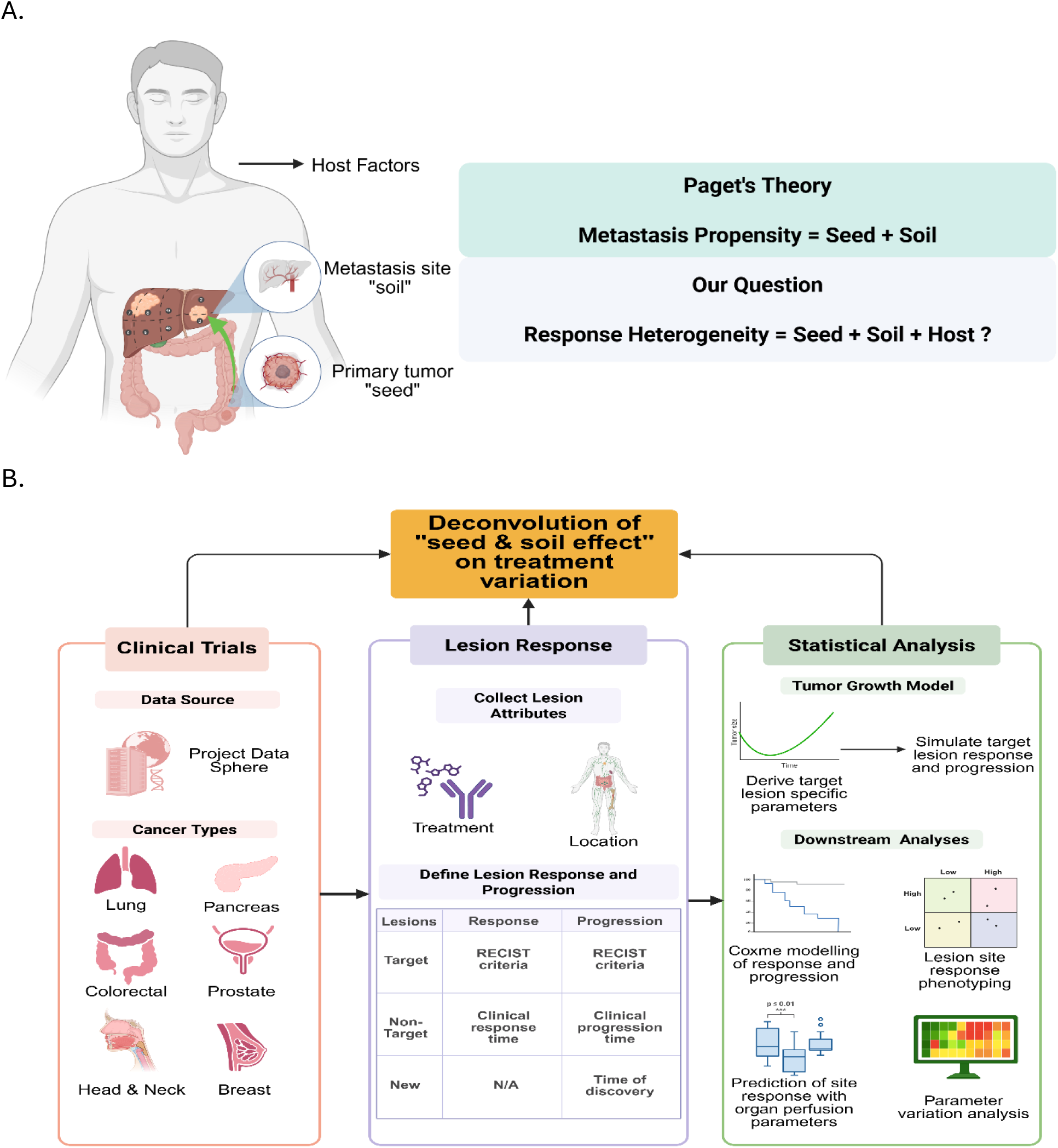
The extended “seed and soil” paradigm (A) and our analytical workflow (B) with statistical modeling approaches used in this pan-cancer analysis.

To rigorously evaluate the extended “seed and soil” paradigm in the context of treatment response, we developed a multi-layered analytical framework (**Figure 1B**) for systematically characterizing lesion-level phenotypic heterogeneity. Building from our previous work conducted by Zhou, et al.(7), lesion growth trajectories were first modeled using a nonlinear mixed-effects (NLME) approach, which enabled estimation of individualized response parameters while accounting for both inter-patient and inter-lesion variability. To isolate the independent contribution of anatomical site, we applied pan-cancer Cox proportional hazards modeling, adjusting for tumor type, treatment modality, and clinical covariates(7). We then projected lesion-level hazard ratios for response and progression onto a two-dimensional map to identify organ-specific response patterns.

Moreover, to provide a physiological tumor type agonistic explanation, we developed the Vascular Perfusion and Leakiness Index (VaPLI), a composite measure integrating organ-specific perfusion and vascular permeability that reflects the vascular distribution potential of therapeutics. In parallel, we considered the immunological context of metastatic sites, such as the immune-privileged status of the brain and the immunologically active environment of the liver, which may influence lesion-level resistance and progression phenotypes, particularly under immunotherapy.

Together, our approach integrates modeling, physiology, and immunology to extend the “seed and soil” concept from a theory of metastatic dissemination to a framework for understanding treatment response heterogeneity. By visualizing and quantifying the response-progression landscape across metastatic organs, we aim to provide actionable insights into the biology of metastatic behavior. Ultimately, this lesion-level perspective may inform more personalized treatment strategies, support the development of site-specific therapeutic interventions, and guide future refinements in clinical response assessment.

## Methods

### Data Acquisition and Processing

Figure 1B illustrates the methodology and data sources used in our analysis. We curated de-identified patient-level datasets from 20 clinical trials across six cancer types: colorectal, non-small cell lung (NSCLC), pancreatic, breast, prostate, and head and neck cancers, via Project Data Sphere (https://www.projectdatasphere.org/). Patients were included when longitudinal observations of target and non-target tumor lesions were available. Target, non-target, and new lesion data were extracted alongside patient-level demographics and survival outcomes. All trial and patient information were summarized in **Table 1** and **2** and Supplementary Table S1-S4.

**Table 1.** – summarizing trials details.

| Study | Cancer(s) | Intervention(s) | Data Arm Availability | Sponsor |
| --- | --- | --- | --- | --- |
| NCT00554229 | Hormone Resistant Prostate Cancer (HRPC) w/ Bone Metastasis | ZD4054 /SOC*<br>Placebo/SOC* | Placebo/SOC* arm | AstraZeneca |
| **NCT00988208** | Metastatic Castrate-Resistant Prostate Cancer | Lenalidomide+ Pred + Doc<br>Pred+Doc | Pred+Doc | Celgene |
| **NCT00519285** | Metastatic Androgen Independent Prostate Cancer | Aflibercept+Pred+Doc<br>Pred+Doc | Pred+Doc | Sanofi |
| **NCT00046527** | Metastatic Breast Cancer | ABI-007<br>Taxol | Taxol | Celgene |
| **NCT00457392** | Advanced/Metastatic Non-Small Cell Lung Cancer | Sunitinib+Erlotinib<br>Erlotinib | Erlotinib | Pfizer |
| **NCT00762034** | Stage IIIB or IV nonsquamous Non-Small Cell Lung Cancer | Pem+Carbo+Beva<br>Pac+Carbo+Beva | Pac+Carbo+Beva | Eli Lilly and Company |
| **NCT00417209** | Advanced Pancreatic Cancer | Larotaxel<br>5-FU/Cape | 5-FU/Cape | Sanofi |
| **NCT01016483** | Metastatic Pancreas Cancer | Pimasertib+Gem<br>Gem | Gemcitabine | EMD Serono |
| **NCT01124786** | Metastatic Pancreas Cancer | CO-1.01<br>Gemcitabine | Gemcitabine | Clovis Oncology, Inc. |
| **NCT00844649** | Metastatic Pancreas Cancer | ABI-007+ Gemcitabine<br>Gemcitabine | Gemcitabine | Celgene |
| **NCT00574275** | Metastatic Pancreas Cancer | Aflibercept+<br>Gemcitabine<br>Gemcitabine | Gemcitabine | Sanofi |
| **NCT00460265** | Recurrent and/or Metastatic Head and Neck Cancer | Pani+Cis+5FU<br>Cis+5FU | Pani+Cis+5FU<br>Cis+5FU | Amgen |
| **NCT00384176** | Metastatic Colorectal Cancer | Cediranib+FOLFOX<br>Beva+FOLFOX | Beva+FOLFOX | AstraZeneca |
| **NCT00457691** | Metastatic Colorectal Cancer | Sunitinib+FOLFIRI<br>FOLFIRI | FOLFIRI | Pfizer |
| **NCT00561470** | Metastatic Colorectal Cancer | Aflibercept+FOLFIRI<br>FOLFIRI | FOLFIRI | Sanofi |
| **NCT00305188** | Metastatic Colorectal Cancer | Xaliproden+FOLFOX<br>FOLFOX | FOLFOX | Sanofi |
| **NCT00272051** | Metastatic Colorectal Cancer | Xaliproden+FOLFOX<br>FOLFOX | FOLFOX | Sanofi |
| **NCT00115765** | Metastatic Colorectal Cancer | Pani+Beva+FOLFOX<br>Pani+Beva+FOLFIRI<br>Beva+FOLFOX<br>Beva+FOLFIRI | Beva+FOLFOX<br>Beva+FOLFIRI | Amgen |
| **NCT00339183** | Metastatic Colorectal Cancer | Pani+FOLFIRI<br>FOLFIRI | FOLFIRI | Amgen |
| **NCT00364013** | Metastatic Colorectal Cancer | Pani+FOLFOX<br>FOLFOX | FOLFOX | Amgen |

Abbreviations: SOC - Standard of Care, Pred - Prednisone, Doc- Docetaxel, Pem – Pemetrexed, Carbo – Carboplatin, Beva – Bevacizumab, Cape – Capecitabine, Pani – Panitumumab

**Table 2.** – pan-cancer patient demographics.

|  |  |
| --- | --- |
| **Total Patients** | **6087** |
| **Lesions per patient (mean (SD))** | 9 (7) |
| **Age, years (mean (SD))** | 61.0 (10.4) |
| **Race (n, %)** |  |
| **American Indian/Alaska Native** | 2 (0.0) |
| **Asian** | 192 (3.2) |
| **Black/African American** | 148 (2.4) |
| **Hispanic/Latino** | 1 (0.0) |
| **Other** | 202 (3.3) |
| **White/Caucasian** | 5511 (90.5) |
| **Not reported** | 31 (0.5) |
| **Cancer type (n, %)** |  |
| **Breast** | 265 (4.4) |
| **Colorectal** | 3157 (51.9) |
| **HNSCC** | 393 (6.5) |
| **NSCLC** | 564 (9.3) |
| **Pancreas** | 612 (10.1) |
| **Prostate** | 1096 (18.0) |
| **Baseline ECOG (n, %)** |  |
| **0** | 935 (15.4) |
| **1** | 540 (8.9) |
| **2** | 8 (0.1) |
| **Not reported** | 4604 (75.6) |
| **Reported progression (n, %)** |  |
| **No** | 2675 (43.9) |
| **Yes** | 3412 (56.1) |
| **Time to progression, days (mean (SD))** | 260.3 (211.3) |
| **Reported death (n, %)** |  |
| **No** | 3121 (51.3) |
| **Yes** | 2966 (48.7) |
| **Time to death, days (mean (SD))** | 449.9 (329.9) |
| **Sex (n, %)** |  |
| **Male** | 3748 (61.6) |
| **Female** | 2072 (34.0) |
| **Not reported** | 267 (4.4) |
| **Treatment group (n, %)** |  |
| **Antibody +/- Chemotherapy** | 2414 (39.7) |
| **Targeted therapy** | 430 (7.1) |
| **Chemotherapy** | 3045 (50.0) |
| **Other** | 198 (3.3) |

All study protocols were approved by the institutional review boards (IRBs) of participating centers, as well as the clinical trial review boards of the sponsor companies listed in **Table 1**. Written informed consent was obtained from all patients before any study-related procedures. All trials included were randomized, double-blind, placebo-controlled Phase III studies.

### Lesion-specific Phenotypic Parameters

Lesion growth and response trajectories were modeled using a modified Stein equation(7):

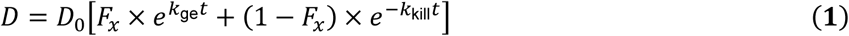

where D is the tumor diameter, D_0_ is the tumor baseline diameter, t is the time. The model has three parameters for estimation: Fx is the fraction of non-responding tumor cells, with 1-Fx as the response depth; k_ge_ is the progression rate and k_kill_ is the regression rate. Figure S1 illustrates the impact of the equation parameters on the lesion growth dynamics curve.

Model fitting was performed using nonlinear mixed-effects (NLME) modeling in Monolix2024R1 with the SAEM algorithm. The M3 method was used for handling measurements below quantification limits (<5 mm)(8,9).

### Tumor Regression and Progression Patterns

Model-estimated tumor shrinkage and growth parameters were used to determine lesion-specific response and progression times per RECIST v1.1 criteria. For target lesions, response was defined as a ≥20% reduction from baseline, and progression as a ≥30% increase from nadir or ≥15 mm absolute growth. For these target lesions, we used NLME model-predicted tumor growth dynamics instead of observed tumor size to determine tumor response and progression because model-based estimates provide a more sensitive and accurate characterization of underlying tumor behavior, accounting for measurement variability, missing data, and inter-individual differences. Non-target lesion responses and progression were annotated using clinical records. The new lesion progression was defined by the date of initial detection.

### Cox Proportional Hazards Modeling

Lesion-level response and progression risks were quantified using Cox proportional hazards models with patient-level random effects. Covariates (e.g., treatment class, prior therapy, BMI, demographics) were tested, and those with p<0.05 were retained. New lesions were considered only in the progression models. Analyses were conducted in R (v4.4.3) using the coxme package.

To assess the independent role of lesion location, pan-cancer Cox models were constructed, adjusting for tumor type, treatment, and significant covariates. Hazard ratios (HRs) for response and progression were plotted in 2D coordinate space to visualize organ-level trends.

### Phenotypic Heterogeneity Across Organ Sites

We quantified anatomical heterogeneity of tumor shrinkage and growth parameters (Fx, kge, kkill) using relative variability and coefficient of variation (CV%).

Relative variability:

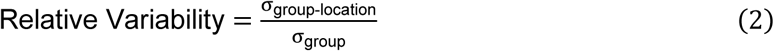

where σ_group-location_ is the standard deviation within a specific group-location, and σ_overall_ is the standard deviation across the group. This normalization enabled identification of locations where parameter fluctuations were disproportionately pronounced relative to overall variability.

Coefficient of Variation (CV%) was also computed for each grouping-location pair as a standardized measure of dispersion. CV% was calculated as the ratio of the standard deviation to the mean parameter value, multiplied by 100, using formula (3).

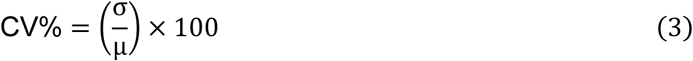

where σ is the standard deviation and µ is the mean within each group-location pair.

### Physiological Predictors: Vascular Permeability and Immune Features

We constructed a Vascular Perfusion and Leakiness Index (VaPLI) to approximate site-specific drug permeability and biodistribution:

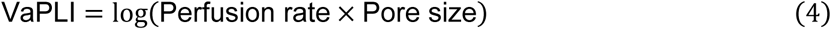

Organ-specific estimates of perfusion rates and vascular permeability were obtained from published literature and pharmacokinetic modeling studies(10–12). These values were standardized and integrated to generate a single continuous index for each anatomical location. Higher VaPLI values reflect greater theoretical drug delivery potential, driven by increased blood flow and/or vascular permeability, whereas lower values indicate limited perfusion and/or more restrictive vasculature. VaPLI scores were then correlated with lesion-level response HRs.

To evaluate immune context, organs were classified as immune-active (e.g., lymph nodes, GI tract, spleen) or immune-tolerant (e.g., brain, liver, bone). Z-score–normalized progression HRs were compared between groups using Wilcoxon rank-sum tests.

## Results

### Lesion-Level Phenotypic Data in this Pan-Cancer Analysis

The breadth of lesion-level data across multiple cancer types supports the generalizability of this pan-cancer analysis. Figure 2A summarizes the inclusion and exclusion criteria used to select 20 metastatic cancer trials (Table 1), encompassing a total of 6,087 patients and 55,220 metastatic lesions. Lesions were approximately evenly distributed between target (n=26,398, 48%) and non-target (n=25,245, 46%) categories, with a smaller subset of new lesions (n=3,577, 6%) also included in the analysis.

**Figure 2.**
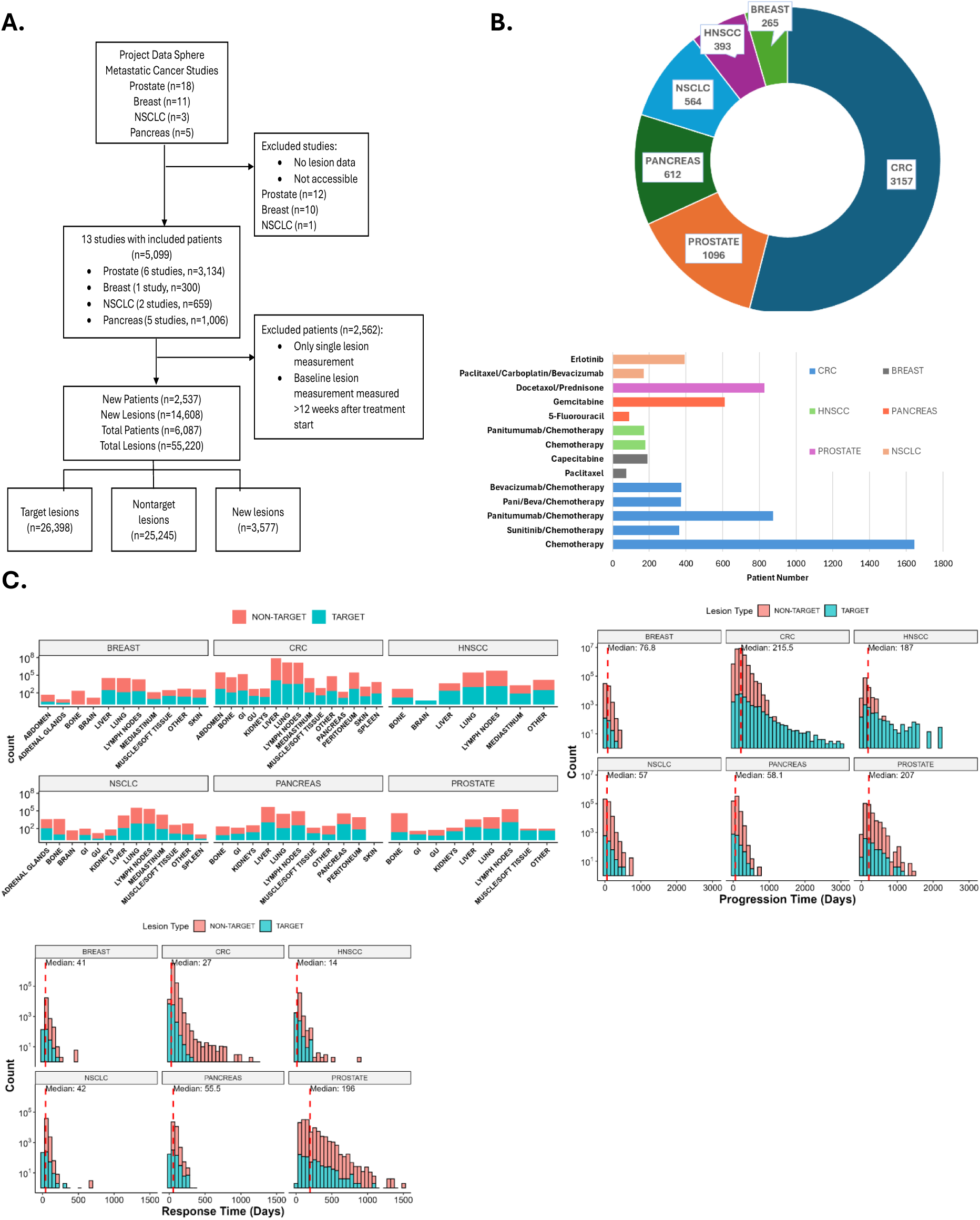
Data source, population characteristics, and lesion features. (A) CONSORT diagram showing inclusion and exclusion criteria for metastatic cancer lesions. The total numbers of patients and lesions include studies from Zhou et al. (B) Number of patients and distribution of treatment types by cancer diagnosis. (C) Distribution of all lesions—new lesions grouped with non-target lesions when applicable— across organ sites, along with corresponding response and progression times.

As shown in Figure 2B, CRC patients comprised the largest share of the dataset (n=3,157), followed by prostate cancer (n=1,096). Chemotherapy was the most common treatment modality, though targeted agents, such as tyrosine kinase inhibitors and monoclonal antibodies, were also represented across the dataset.

Figure 2C presents lesion distribution by cancer type, illustrating that liver, lung, bone, and lymph nodes were the most frequent metastatic sites. Brain metastases were particularly prevalent in NSCLC and breast cancer cohorts compared to others. The lower panel of Figure 2C shows variability in clinical response and progression dynamics by cancer type. CRC and breast cancer patients demonstrated the shortest median times to response, while prostate cancer patients had the longest. In contrast, pancreatic and NSCLC patients exhibited more rapid disease progression, whereas prostate and CRC patients experienced longer progression-free intervals. The heterogeneous distribution of target and non-target lesions highlights the importance of evaluating organ– and cancer-specific patterns. The response probability and duration of response for each cancer type, across different treatment groups, are shown in Figure S1.

### Lesion Regression and Progression Reflects Strong Seed-Soil Interplay

Lesion-level tumor growth modeling yielded estimates for three key parameters: kkill (tumor regression rate), kge (tumor progression rate), and Fx (fraction of treatment-resistant tumor cells). The goodness of fitting (Figure S2) and representative individual lesions (Figure S3) both show adequate model performance. Figure 3A illustrates the distribution of these parameters in two representative cancer types, prostate cancer and head and neck squamous cell carcinoma (HNSCC), across various metastatic sites. In prostate cancer, bone lesions exhibited statistically significant differences compared to other sites, including the liver (kkill), gastrointestinal tract (Fx), and lymph nodes (kge), suggesting that the local microenvironment plays a critical role in modulating tumor growth and treatment response in prostate cancer. Similarly, liver metastases in HNSCC displayed distinct parameter profiles, consistent with both increased treatment sensitivity and potentially accelerated tumor progression. Parameter estimates for other cancer types were summarized in Figure S4.

**Figure 3.**
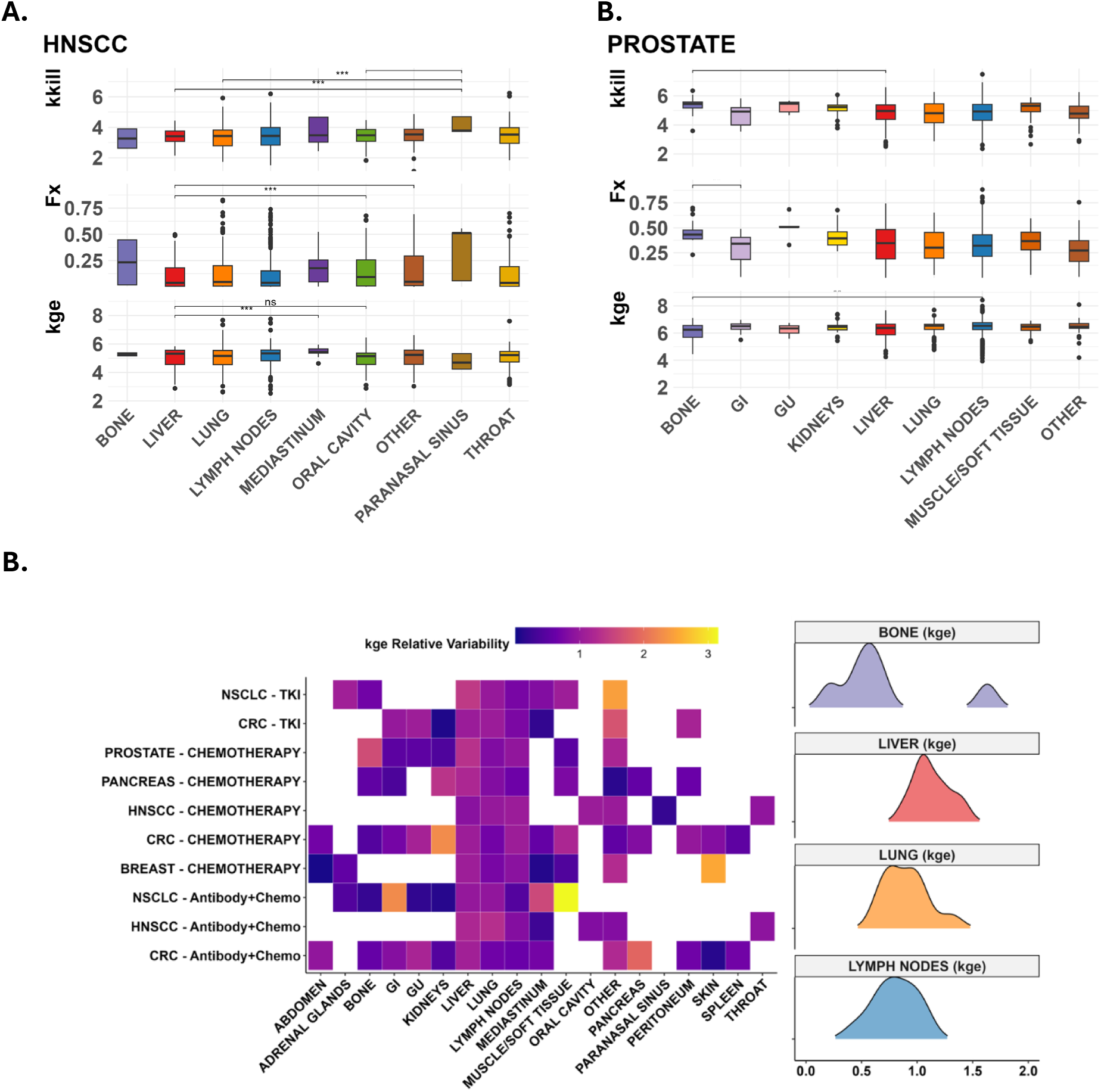
Variability in tumor regression and progression across metastatic sites (‘soils’), highlighting trends and differences by cancer type (‘seeds’) and treatment combinations. **A** Statistically significant parameter differences across anatomical locations (soil) with p<0.05 by ANOVA. The liver shows statistically significant differences in cross-wise comparisons for head & neck cancer while the bone lesions show variation in prostate cancer using Wilcoxon test and correcting using FDR (p<0.05). **B** The heatmaps show relative variability of each parameter by cancer-treatment group in a organ/site; a value greater than 1 means that there is greater intra-site variability at than at the cancer level, indicating heterogeneity due to host and unknown factors.

Figure 3B highlights the variability of these growth and response parameters across cancer-treatment-site combinations. Summary distributions for select metastatic sites are shown on the right. A strong seed–soil interaction is evident in the tumor growth (kge) parameter, further modulated by treatment type. The kge parameter exhibited greater variability in the kidneys, muscle/soft tissue, and gastrointestinal tissue, while the bone. As shown In Figure S5, the Fx parameter remained relatively stable across most anatomical sites, particularly in the lung and lymph nodes. Notably, kkill showed pronounced heterogeneity in liver lesions, underscoring the potential influence of host-specific factors and unmeasured biological modifiers in this organ.

Liver lesions demonstrated high variability across all three parameters, in contrast to bone lesions, which showed more consistent profiles for kkill and kge, reflecting a potentially more uniform microenvironment. These findings emphasize the importance of considering not only the tumor lineage (“seed”) and metastatic site (“soil”) but also the treatment modality and host-specific biological context when evaluating lesion-level response dynamics.

### Lesion-Level Treatment Response Reflects Seed–Soil–Treatment Interactions

Lesion response and progression behaviors were systematically analyzed to uncover spatial patterns of therapeutic sensitivity and resistance. To ensure comprehensive evaluation, both target and non-target lesions were included. Emphasis was placed on common metastatic sites with high lesion counts to enable robust statistical comparisons. Figure 4 summarizes lesion-level response probabilities (defined as >20% tumor shrinkage), hazard ratios for response and progression, response durations, and correlations between lesion response and progression.

**Figure 4.**
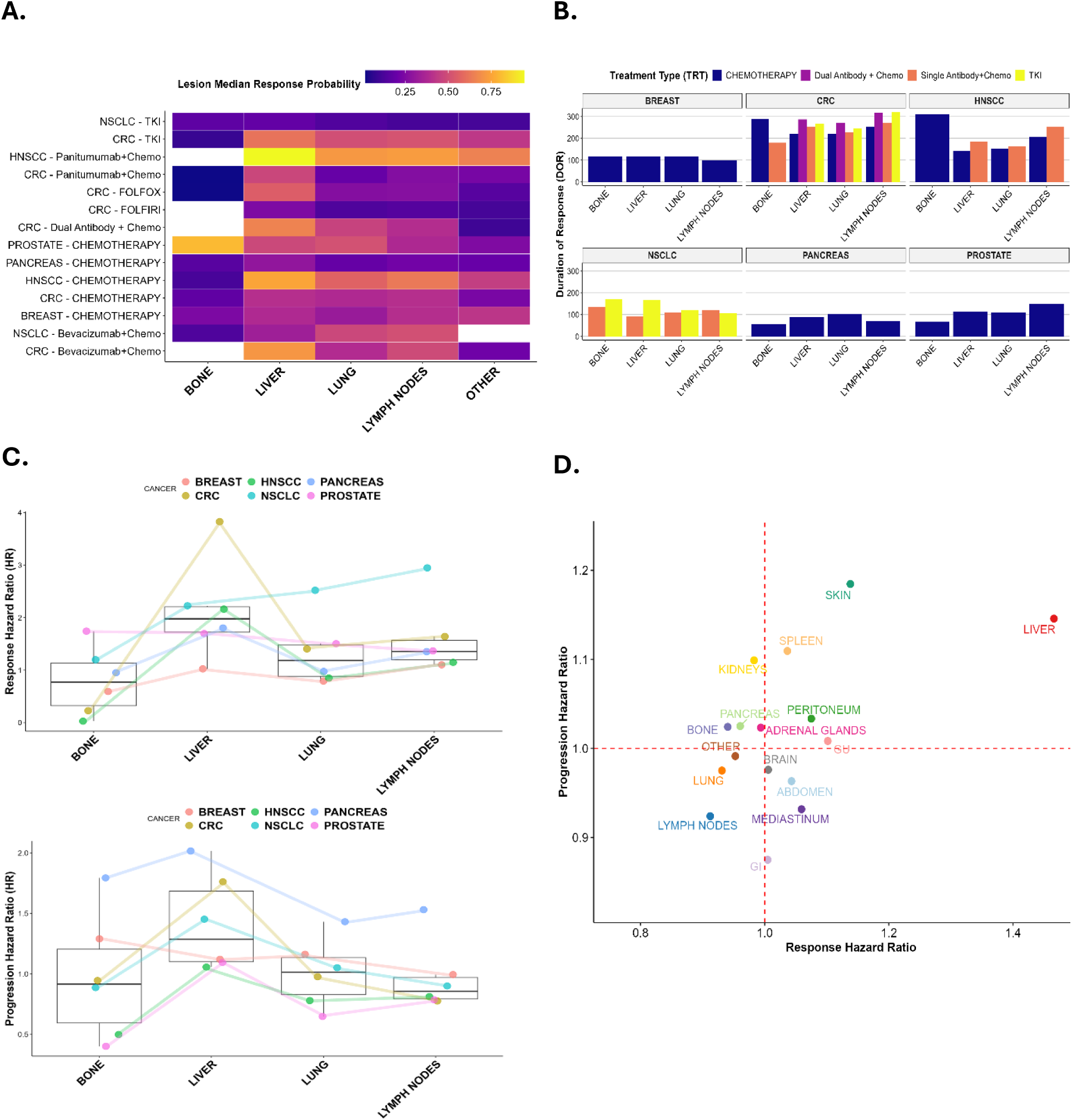
Both “soil” and “seed” influence lesion response rate, durability of response, and progression hazard across different treatments. **A** The heatmap displays the median lesion probability of response (displayed as decimal value) across soil organs/sites within cancer-treatment groups. **B** The bar chart shows the variability in lesion duration of response is shown across sites within cancer patients grouped by treatment type. **C** Plots of the common organ/sites across cancer types analyzed using coxme interaction analysis show trends both intra-cancer and intra-site. **D** Cartesian grid of response rates and progression hazard ratios for each soil organ/site, controlling for cancer type and treatment.

Figure 4A displays response probabilities stratified by cancer type, treatment modality, and lesion location. Liver metastases exhibited the highest, but also the most heterogeneous, response probabilities across cancer types. In contrast, lesions in lymph nodes and lungs showed more moderate and consistent response rates. Notably, prostate cancer lesions in bone demonstrated significantly higher response probabilities compared to bone lesions from other cancers, suggesting that tumor–host interactions within the osseous microenvironment may enhance treatment sensitivity.

Figure 4B illustrates response duration patterns across cancer types and lesion sites. Breast and pancreatic cancers showed relatively uniform response durations across metastatic locations, whereas colorectal cancer (CRC) and non-small-cell lung cancer (NSCLC) exhibited substantial heterogeneity. Targeted therapies, including monoclonal antibodies and tyrosine kinase inhibitors (TKIs), were associated with longer progression-free survival in CRC, NSCLC, and head-and-neck squamous cell carcinoma (HNSCC), particularly in select metastatic sites.

Figure 4C presents hazard ratios for response and progression, modeled by lesion location and cancer type while adjusting for treatment effects. Liver lesions consistently showed elevated hazard ratios for both response and progression, reinforcing prior observations of their distinct clinical behavior. Trend lines across cancer types highlight how tumor-intrinsic (“seed”) factors contribute to site-specific dynamics, providing additional support for the extended “seed and soil” paradigm. Full model results are available in the Supplementary Information.

To isolate the effects of anatomical site independent of cancer type and therapy, Figure 4D shows adjusted lesion-specific hazard ratios by metastatic location. Liver metastases continued to demonstrate both high response and rapid progression, while lymph node lesions exhibited lower response hazards and reduced progression risks, suggesting a more controlled disease trajectory in these sites.

Collectively, these findings underscore the multifactorial nature of lesion-level treatment response. The convergence of tumor lineage (seed), anatomical location (soil), and treatment modality drives variability in therapeutic outcomes. These insights may inform future precision oncology strategies and support the development of site-aware therapeutic decision-making frameworks.

### Two “Soil” Physiological Features Predict Lesion-Level Treatment Response

We further analyzed the microenvironmental factors (“soil” properties) that influence lesion-level treatment responses. Specifically, we evaluated the role of vascular perfusion, vascular permeability, and immune landscape in shaping therapeutic efficacy across metastatic sites. The organ-specific capillary fenestration sizes and perfusion rates used for VaPLI calculation were summarized in Figure S6.

In Figure 5A, the VaPLI was used to quantify the theoretical drug delivery potential of each organ site. The spleen and liver showed the highest VaPLI values, indicating enhanced exposure to systemic anticancer therapies. In contrast, bone, brain, and muscle tissues exhibited the lowest VaPLI scores, reflecting restricted vascular delivery.

**Figure 5.**
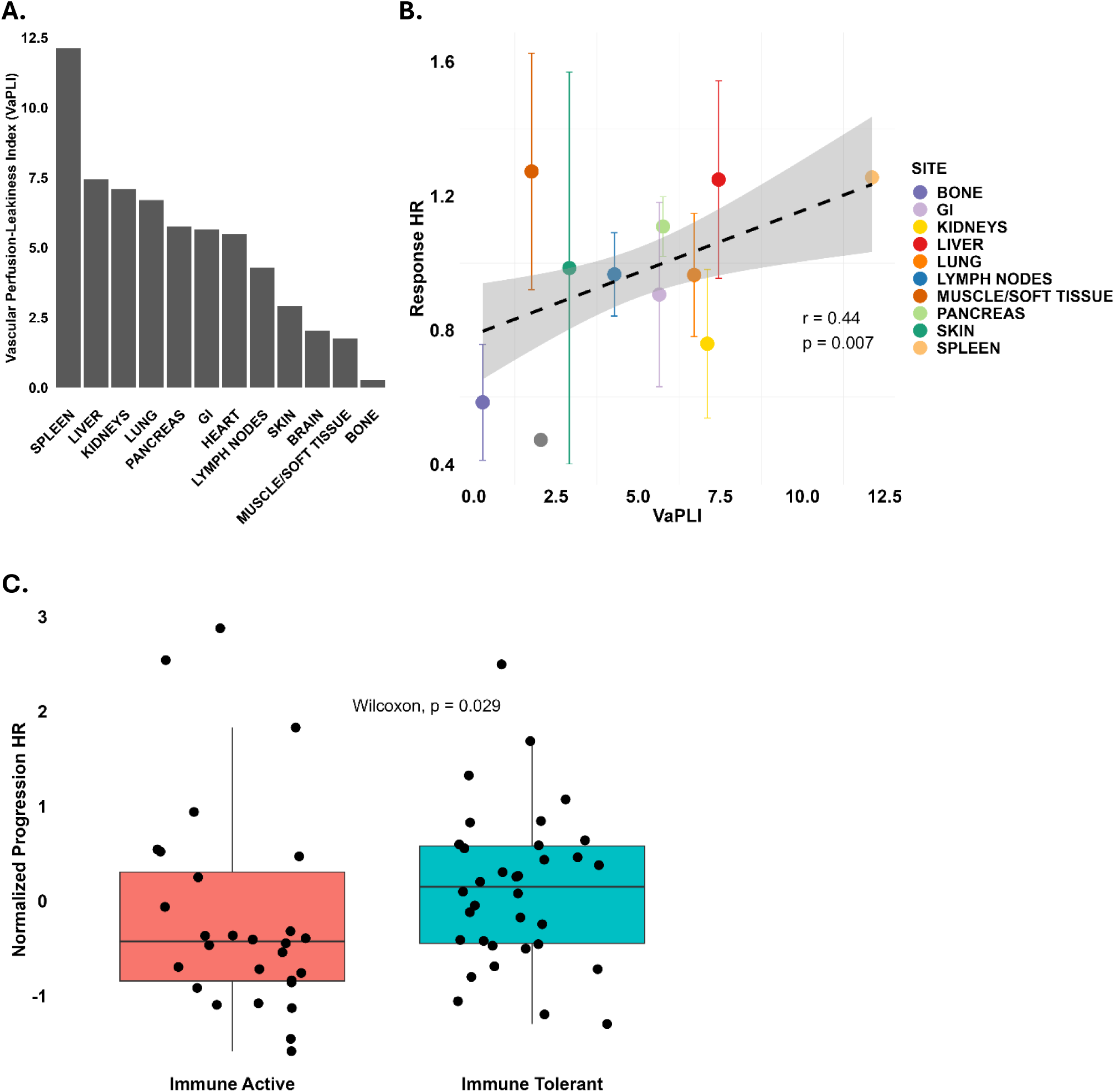
Vascular perfusion and leakiness (A) predict response trends across “soils” (B) and the immune landscape of anatomical organs predicts progression risk (C). **A** The Vascular Perfusion Leakiness Index (VaPLI) values for organ/sites are shown. **B** The x-axis plots the VaPLI values corresponding to the organ/site (color). The y-axis plots the response rate for each cancer-site pairing, controlled for treatment. Error bars represent the standard error in mean HRs across cancer types. A statistically significant, moderate relationship is seen between increasing VaPLI values and organ response rates. **C** This boxplot shows a statistically significant difference (p < 0.05, Wilcoxon rank-sum test) in progression hazard ratios between immune-active and immune-tolerant organ groups, following Z-score normalization of the hazard ratios, within each cancer type. Immune-tolerant sites: brain, liver, muscle/soft tissue, kidneys, pancreas, bone. Immune-active sites: spleen, skin, lung, lymph nodes, GI tract.

As shown in Figure 5B, there is a correlation analysis between VaPLI and lesion-specific response hazard ratios. A statistically significant moderate positive correlation was observed, suggesting that higher perfusion and vascular permeability are associated with greater likelihood of therapeutic response. These findings highlight the critical role of physiological accessibility in shaping lesion-level outcomes and reinforce the importance of incorporating organ-specific vascular characteristics into predictive models of treatment efficacy.

The immune landscape of metastatic sites appears to influence progression risk. As shown in Figure 5C, despite variability across cancer types and treatment modalities, tissues characterized by immune tolerance, such as the liver exhibit, on average, higher progression hazards compared to tissues with active immune surveillance.

## Discussion

Our analysis of 55,220 metastatic lesions from 6,087 patients across 20 clinical trials highlights the critical and multifaceted influence of both tumor-intrinsic (“seed”) and organ-specific (“soil”) factors on metastatic progression and therapeutic response. By systematically modeling lesion-level tumor growth and response dynamics across diverse cancer types, treatments, and anatomical sites, we extend the classical “seed and soil” paradigm beyond metastatic colonization to include treatment sensitivity and progression behavior at the lesion level.

A central insight from this extended framework is that the organ microenvironment significantly modulates therapeutic response, independent of tumor lineage or drug class. For instance, liver metastases consistently demonstrated high initial response rates but were also associated with rapid progression, suggesting an environment conducive to drug delivery but prone to resistance emergence. In contrast, bone lesions, particularly in prostate cancer, showed more stable regression patterns, yet exhibited variability in progression across cancer types, indicating that both “seed” (tumor) and “soil” (microenvironment) interactions jointly govern lesion treatment response behaviors.

This site-specific variability was further supported by mechanistic correlates. Organ-level vascular characteristics, summarized by the Vascular Perfusion and Leakiness Index (VaPLI), were positively associated with response hazard ratios. Tissues with high perfusion and permeability, such as the liver and spleen, likely enable rapid drug penetration and early tumor regression. However, these same features may facilitate clonal expansion and therapy resistance due to high cellular turnover(13). Additionally, we evaluated how immune contexture influences progression risk across organ sites. Organs were classified as immune-active or immune-tolerant based on literature describing tissue-specific immune surveillance and T cell activity. Using the Z-score normalized hazard ratios from multivariable Cox mixed-effects models, we observed significantly higher progression risk in immune-tolerant organs(14–19) — including the brain, bone, liver, pancreas, and kidneys — compared to immune-active sites such as the lung, skin, and gastrointestinal tract(20–24). This observation, confirmed by Wilcoxon rank-sum testing, supports the notion that immune-privileged environments may impair durable treatment efficacy despite initial response, aligning with prior studies implicating local immunosuppression as a barrier to sustained therapeutic control(25).

By expanding the “seed and soil” paradigm to encompass therapeutic outcomes, our findings call attention to a critical but underappreciated source of heterogeneity in cancer care: the spatial distribution of metastatic disease. The same treatment may yield vastly different outcomes across lesions within a patient, not solely due to tumor heterogeneity but also because of local physiological and immunological conditions.

These insights carry important implications for clinical trial design and drug development. Conventional endpoints such as RECIST, which rely on aggregate measures or a limited subset of target lesions, may obscure organ-specific treatment failures or successes. Future trials may benefit from stratifying response analyses by metastatic site or incorporating lesion-level endpoints that reflect the biology of individual metastatic niches.

Therapeutic planning should also account for the anatomical distribution of metastases. Patients with liver-dominant disease may respond rapidly but require intensified or combination regimens to prevent early progression. Conversely, bone-predominant disease, particularly in prostate cancer, may respond more slowly but durably, supporting distinct therapeutic pacing. Moreover, drug delivery strategies that enhance vascular access or modulate the immune contexture of specific tissues could improve outcomes in historically refractory metastatic sites such as the brain and bone.

Several limitations of this study should be acknowledged. First, although the dataset encompasses a broad spectrum of cancer types and therapeutic modalities, variability in imaging protocols, lesion measurement frequency, and clinical assessment criteria across trials may introduce noise or bias into lesion-specific parameter estimates. Second, the tumor growth model employed reflects a simplified dynamic and may not fully capture complex biological processes such as immune-mediated effects, clonal evolution, or spatial heterogeneity within lesions. Third, our modeling of vascular exposure relied on the Vascular Perfusion and Leakiness Index (VaPLI), derived from published physiological data, rather than direct perfusion or drug distribution measurements in individual patients—potentially limiting mechanistic resolution. Similarly, immune landscape assessments were based on known organ-level immunological characteristics, not patient-specific immune profiling, which restricts inference regarding immune-modulated progression(25–27). Finally, while our lesion-level analysis improves spatial granularity, it does not explicitly address the collective behavior of multiple lesions within a patient or how inter-lesional dynamics influence overall clinical outcomes, an important area for future research.

Together, these insights support an extended “seed and soil” paradigm—one that incorporates not only the metastatic potential of tumor cells and the receptivity of the microenvironment but also their interactions with systemic therapy and host physiology. This framework offers a novel lens through which to design site-aware treatment strategies, develop location-specific therapeutics, and refine clinical trial endpoints, ultimately aiming for more personalized and durable responses in metastatic cancer.

## Additional Information

## Acknowledgements

The authors have no acknowledgments.

## Authors’ contributions

ID conducted analyses, figure creation, and led manuscript writing. JZ helped guide analysis, modeling, and helped write the manuscript. QF helped with statistical methods and helped review manuscript. YC served as advisor of project and helped write the manuscript.

## Ethics approval and consent to participate

All study protocols were approved by the institutional review boards (IRBs) of participating centers, as well as the clinical trial review boards of the sponsor companies listed in Table 1. Written informed consent was obtained from all patients before any study-related procedures. All trials included were randomized, double-blind, placebo-controlled Phase III studies. All studies were performed in accordance with the Declaration of Helsinki.

## Consent for publication

N/A

## Data availability

Clinical trial data are available publicly on Project Data Sphere. Other datasets for this manuscript are available as part of the Supplementary Information.

## Competing interests

The authors declare no conflict of interest.

## Funding information

National Institutes of Health (NIH R35 GM152449 to Y.C.) and Dilley Ian is supported by the National Institute of General Medical Sciences of the National Institutes of Health under award number T32GM086330 (University of North Carolina at Chapel Hill-Duke Collaborative Clinical Pharmacology T32 Postdoctoral Training Program).

## Notes

### Competing Interest Statement

The authors have declared no competing interest.

### Author Declarations

Data availability: The study used (or will use) ONLY openly available human data that were originally located at:(https://www.projectdatasphere.org/).

