## Supplementary for "A Pan-Cancer Analysis of Lesion-Level Treatment Response to Extend the ‘Seed and Soil’ Paradigm"

**Supplementary Materials**

**Extending the 'Seed and Soil' Paradigm: A Pan-Cancer Analysis of Lesion-Specific Therapeutic Response and Progression**

Dilley Ian^1^, Zhou Jiawei^1,2^, Li Quefeng^3^, Cao Yanguang^1,2*^

1 Division of Pharmacotherapy and Experimental Therapeutics, UNC Eshelman School of Pharmacy, University of North Carolina at Chapel Hill, Chapel Hill, NC, 27599, USA.

2 Lineberger Comprehensive Cancer Center, School of Medicine, University of North Carolina at Chapel Hill, Chapel Hill, NC 27599, USA.

3 Department of Biostatistics, Gillings School of Global Public Health, University of North Carolina at Chapel Hill, Chapel Hill, NC, 27599, USA.

Supplement Table 1 – Prostate cancer patient & lesion baseline information

| **Patient demographics** | **DOCETAXEL/ PREDNISONE** | **PREDNISONE/ PLACEBO** |
| --- | --- | --- |
| **n** | 845 | 251 |
| **PSA (mean (SD))** | 187.47 (351.05) | 324.35 (586.91) |
| **ALBUMIN (mean (SD))** | 4.13 (0.42) | 4.15 (0.34) |
| **ALT (mean (SD))** | 24.15 (15.33) | 21.55 (14.55) |
| **AST (mean (SD))** | 27.73 (12.64) | 27.88 (15.73) |
| **ALKALINE_PHOSPHATASE  (mean (SD))** | 264.03 (369.47) | 214.48 (353.58) |
| **CREATININE  (mean (SD))** | 0.97 (0.21) | 0.83 (0.25) |
| **TESTOSTERONE  (mean (SD))** | 17.43 (20.47) | 292.01 (601.34) |
| **AGE (mean (SD))** | 67.27 (7.71) | 67.76 (7.58) |
| **ECOG (%)** |  |  |
| **0** | 688 (81.4) | 130 (52.0) |
| **1** | 149 (17.6) | 120 (48.0) |
| **2** | 8 (0.9) | 0 (0.0) |
| **RACE (%)** |  |  |
| **ASIAN** | 35 (4.3) | 27 (10.8) |
| **BLACK** | 27 (3.3) | 8 (3.2) |
| **OTHER** | 16 (2.0) | 30 (12.0) |
| **WHITE** | 738 (90.4) | 186 (74.1) |
| **RESPONSE = 1 (%)** | 274 (32.7) | 0 (0.0) |
| **RELAPSE = 1 (%)** | 295 (35.2) | 58 (57.4) |
| **STUDY (%)** |  |  |
| **6650** | 0 (0.0) | 251 (100.0) |
| **8208** | 267 (31.6) | 0 (0.0) |
| **9285** | 578 (68.4) | 0 (0.0) |

| **Lesion data** | **NON-TARGET** | **TARGET** |
| --- | --- | --- |
| **n** | 2902 | 2219 |
| **RESPONSE = 1 (%)** | 408 (14.1) | 962 (56.3) |
| **RELAPSE = 1 (%)** | 1652 (56.9) | 538 (31.5) |
| **SITE (%)** |  |  |
| **ABDOMEN** | 3 (0.1) | 8 (0.4) |
| **ADRENAL** | 1 (0.0) | 33 (1.5) |
| **BLADDER** | 9 (0.3) | 6 (0.3) |
| **BONE** | 2363 (81.4) | 24 (1.1) |
| **GI** | 1 (0.0) | 2 (0.1) |
| **KIDNEYS** | 3 (0.1) | 2 (0.1) |
| **LIVER** | 28 (1.0) | 222 (10.0) |
| **LUNGS** | 137 (4.7) | 126 (5.7) |
| **LYMPH NODES** | 258 (8.9) | 1581 (71.6) |
| **MUSCLE/SOFT TISSUE** | 2 (0.1) | 46 (2.1) |
| **OTHER** | 23 (0.8) | 90 (4.1) |
| **PLEURA** | 11 (0.4) | 1 (0.0) |
| **PROSTATE** | 63 (2.2) | 68 (3.1) |
| **TRT = PREDNISONE/ PLACEBO (%)** | 853 (29.4) | 499 (22.5) |
| **STUDY (%)** |  |  |
| **6650** | 853 (29.4) | 499 (22.5) |
| **8208** | 0 (0.0) | 607 (27.4) |
| **9285** | 2049 (70.6) | 1113 (50.2) |

| **Lesion data** | **NON-TARGET** | **TARGET** |
| --- | --- | --- |
| **n** | 1945 | 1665 |
| **RESPONSE = 1 (%)** | 223 (11.5) | 531 (31.9) |
| **RELAPSE = 1 (%)** | 1280 (65.8) | 929 (55.8) |
| **SITE (%)** |  |  |
| **Adrenal Glands** | 37 (1.9) | 102 (6.1) |
| **Bone** | 446 (22.9) | 9 (0.5) |
| **Brain** | 50 (2.6) | 0 (0.0) |
| **GI** | 11 (0.6) | 8 (0.5) |
| **GU** | 8 (0.4) | 2 (0.1) |
| **Kidney** | 11 (0.6) | 6 (0.4) |
| **Liver** | 129 (6.6) | 129 (7.7) |
| **Lung** | 519 (26.7) | 672 (40.4) |
| **Lymph Nodes** | 339 (17.4) | 624 (37.5) |
| **Mediastinum** | 277 (14.2) | 84 (5.0) |
| **Muscle** | 6 (0.3) | 3 (0.2) |
| **Other** | 82 (4.2) | 9 (0.5) |
| **Retroperitoneum** | 2 (0.1) | 4 (0.2) |
| **Soft Tissue** | 23 (1.2) | 11 (0.7) |
| **Spleen** | 5 (0.3) | 2 (0.1) |
| **TRT = Erlotinib (%)** | 1251 (64.3) | 1121 (67.3) |

Supplement Table 2 – NSCLC cancer patient & lesion baseline information

| **Patient demographics** | **Paclitaxel, Carboplatin  and Bevacizumab** | **Erlotinib** |
| --- | --- | --- |
| **n** | 171 | 393 |
| **AGE (mean (SD))** | 64.38 (8.86) | 60.54 (9.19) |
| **BMI (mean (SD))** | 26.04 (5.16) | 25.91 (5.23) |
| **WT (mean (SD))** | 76.23 (16.67) | 72.98 (16.56) |
| **END_OF_STUDY (mean (SD))** | 146.14 (110.00) | 147.02 (133.30) |
| **PFSDY (mean (SD))** | 149.90 (117.25) | 129.32 (131.54) |
| **SEX = MALE (%)** | 101 (59.1) | 240 (61.1) |
| **RACE (%)** |  |  |
| **American Indian/ Alaska Native** | 2 (1.2) | 0 (0.0) |
| **Asian** | 0 (0.0) | 41 (10.4) |
| **Black** | 18 (10.5) | 4 (1.0) |
| **Other** | 0 (0.0) | 10 (2.5) |
| **White** | 151 (88.3) | 338 (86.0) |

| **Lesion data** | **NON-TARGET** | **TARGET** |
| --- | --- | --- |
| **n** | 841 | 744 |
| **RESPONSE = 1 (%)** | 192 (25.9) | 334 (44.9) |
| **RELAPSE = 1 (%)** | 675 (91.0) | 236 (31.7) |
| **SITE (%)** |  |  |
| **Abdomen** | 16 (1.9) | 3 (0.4) |
| **Adrenal Glands** | 4 (0.5) | 2 (0.3) |
| **Bone** | 226 (26.9) | 0 (0.0) |
| **Brain** | 15 (1.8) | 0 (0.0) |
| **Breast** | 27 (3.2) | 37 (5.0) |
| **Liver** | 122 (14.5) | 298 (40.1) |
| **Lung** | 255 (30.3) | 132 (17.7) |
| **Lymph Nodes** | 101 (12.0) | 196 (26.3) |
| **Mediastinum** | 15 (1.8) | 9 (1.2) |
| **Other** | 23 (2.7) | 23 (3.1) |
| **Skin** | 27 (3.2) | 15 (2.0) |
| **Soft Tissue** | 10 (1.2) | 29 (3.9) |

Supplement Table 3 – Breast cancer patient & lesion baseline information

| **Patient demographics** | **CAPECITABINE** | **TAXOL** |
| --- | --- | --- |
| **n** | 190 | 75 |
| **RACE (%)** |  |  |
| **black** | 0 (0.0) | 1 (1.3) |
| **Other** | 47 (24.7) | 0 (0.0) |
| **white** | 143 (75.3) | 74 (98.7) |
| **ECOG (%)** |  |  |
| **0** | 112 (59.6) | 20 (26.7) |
| **1** | 64 (34.0) | 53 (70.7) |
| **2** | 12 (6.4) | 2 (2.7) |
| **PRSURG = 1 (%)** | 9 (4.7) | 49 (65.3) |
| **PRRAD = 1 (%)** | 163 (85.8) | 50 (66.7) |
| **PRHOR = 1 (%)** | 0 (NaN) | 38 (50.7) |
| **PRCHEM = 1 (%)** | 0 (NaN) | 65 (87.8) |
| **ER = 1 (%)** | 109 (59.6) | 13 (31.7) |
| **PR = 1 (%)** | 78 (45.9) | 8 (25.0) |

| **Lesion data** | **NON-TARGET** | **TARGET** |
| --- | --- | --- |
| **n** | 1530 | 2762 |
| **NEWLES = 1 (%)** | 21 (1.4) | 0 (0.0) |
| **RESPONSE = 1 (%)** | 179 (11.7) | 984 (35.6) |
| **RELAPSE = 1 (%)** | 625 (98.1) | 764 (27.7) |
| **SITE (%)** |  |  |
| **OTHER** | 41 (2.7) | 15 (0.5) |
| **BONE** | 42 (2.7) | 7 (0.3) |
| **GI TRACT** | 13 (0.8) | 15 (0.5) |
| **KIDNEYS** | 16 (1.0) | 26 (0.9) |
| **LIVER** | 497 (32.5) | 1422 (51.5) |
| **LUNGS** | 336 (22.0) | 140 (5.1) |
| **LYMPH NODES** | 302 (19.7) | 408 (14.8) |
| **MUSCLE/ SOFT TISSUE** | 13 (0.8) | 23 (0.8) |
| **PANCREAS** | 103 (6.7) | 631 (22.8) |
| **PERITONEUM** | 167 (10.9) | 75 (2.7) |
| **TRT = Gemcitabine** | 1250 (81.7) | 2232 (80.8) |

Supplement Table 4 – Pancreatic cancer patient & lesion baseline information

| **Patient demographics** | **5-FU** | **Gemcitabine** |
| --- | --- | --- |
| **n** | 92 | 611 |
| **AGE (mean (SD))** | 58.72 (9.39) | 62.84 (9.05) |
| **ALP (mean (SD))** | 202.31 (180.55) | 204.51 (193.81) |
| **AST (mean (SD))** | 28.85 (16.30) | 33.54 (20.66) |
| **SEX = Male (%)** | 50 (54.3) | 346 (56.6) |
| **RACE (%)** |  |  |
| **ASIAN** | 0 (0.0) | 8 (1.3) |
| **BLACK** | 0 (0.0) | 13 (2.1) |
| **CAUCASIAN / WHITE** | 84 (91.3) | 579 (94.9) |
| **OTHER** | 8 (8.7) | 10 (1.6) |

Supplement Figure 1

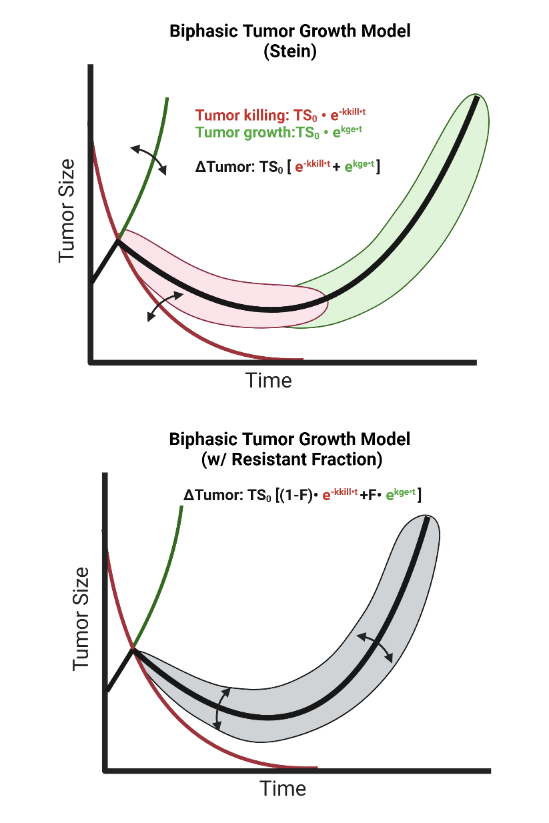

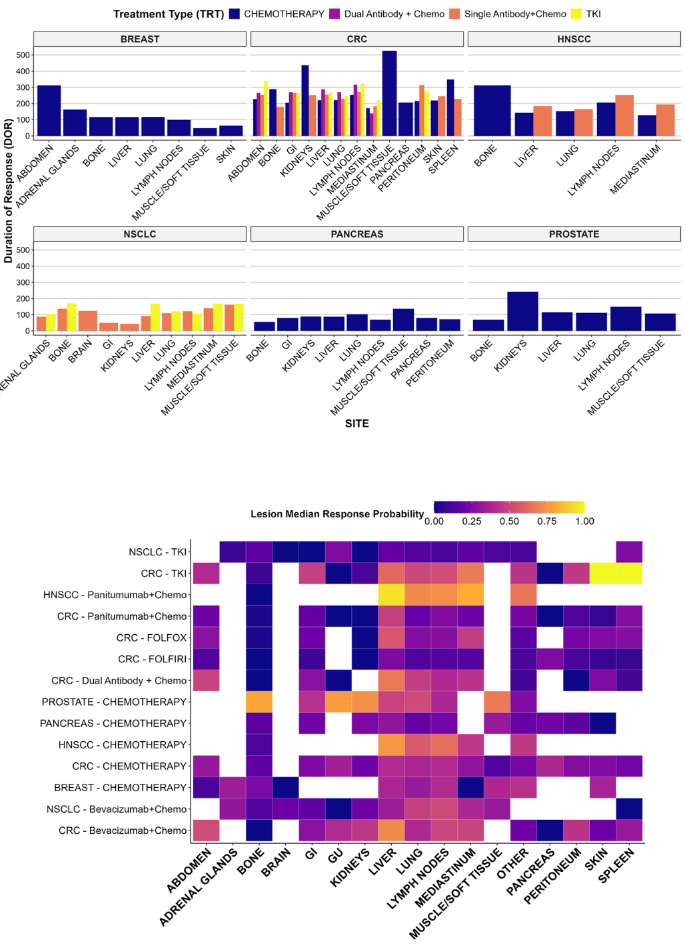

Left panel illustrates impact of the various TGI parameters on the lesion growth trajectories. Right panel displays duration of response distribution within cancer types across various lesion locations, as well as the response probability heatmap for various lesion locations.

Supplement Figure 2

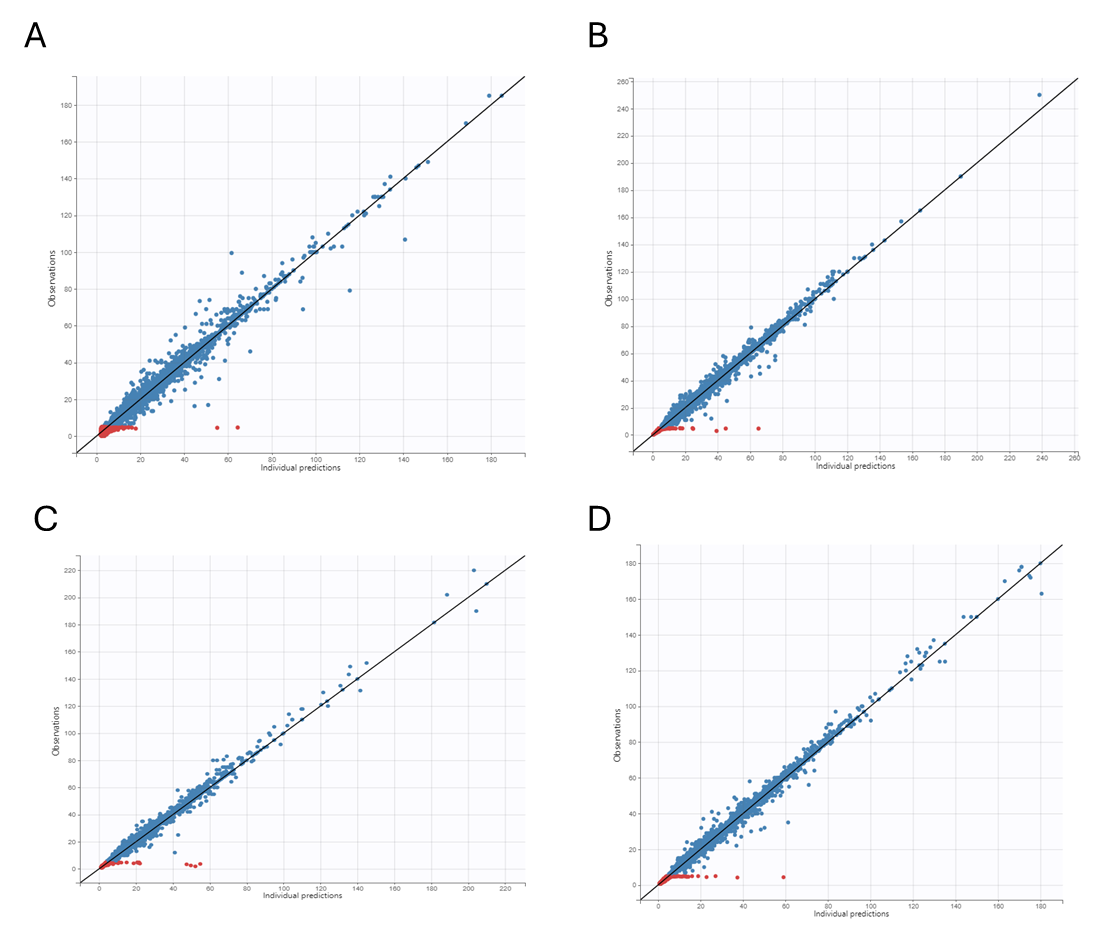

Goodness-of-fit plots for lesion growth predictions vs. observations for A) prostate cancer, B) NSCLC, C) breast cancer, and D) pancreatic cancer. Alignment along line of unity indicates appropriate fit across lesion sizes. Red dots indicate BLQ data (<5mm).

Supplement Figure 3

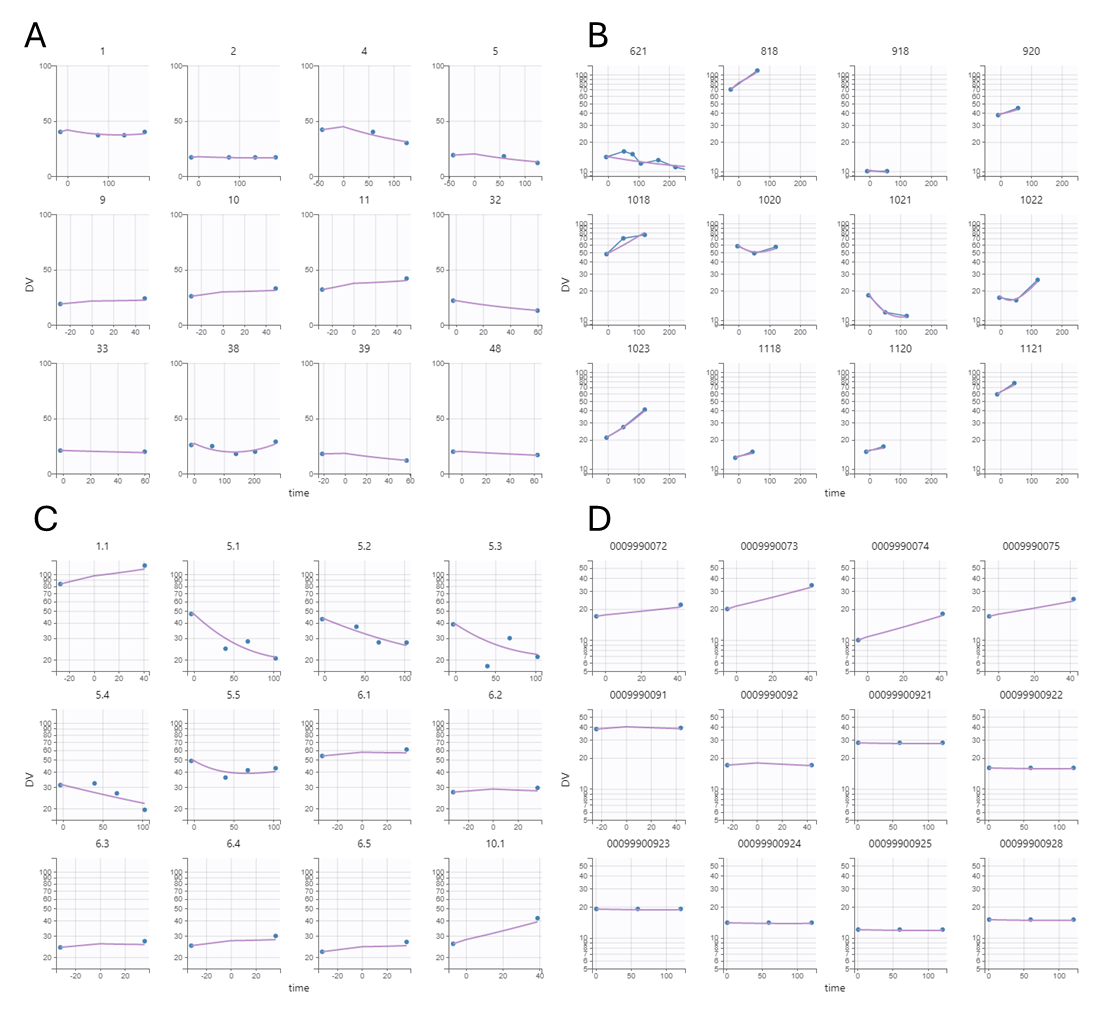
Individual lesion growth curve examples fit plots for A) prostate cancer, B) NSCLC, C) breast cancer, and D) pancreatic cancer. Blue dots represent observations, and purple line displays simulated growth trajectory for the individual lesion.

Supplement Figure 4

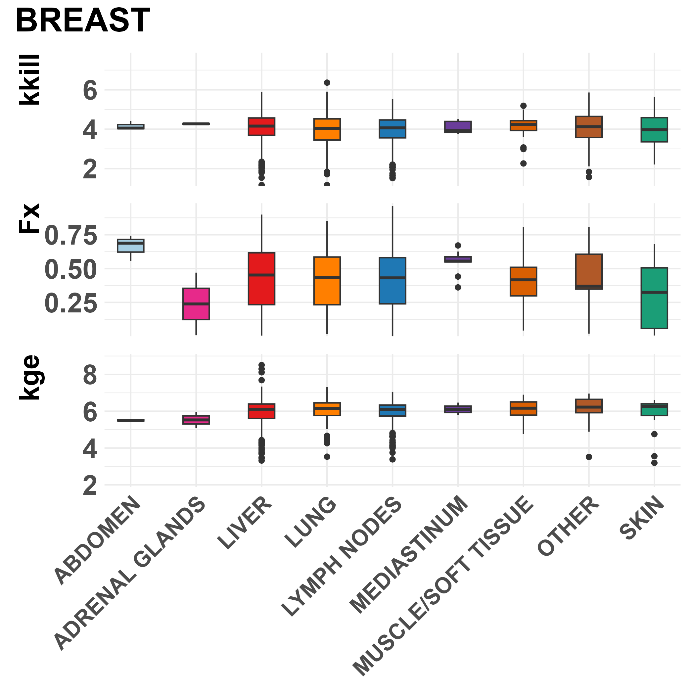

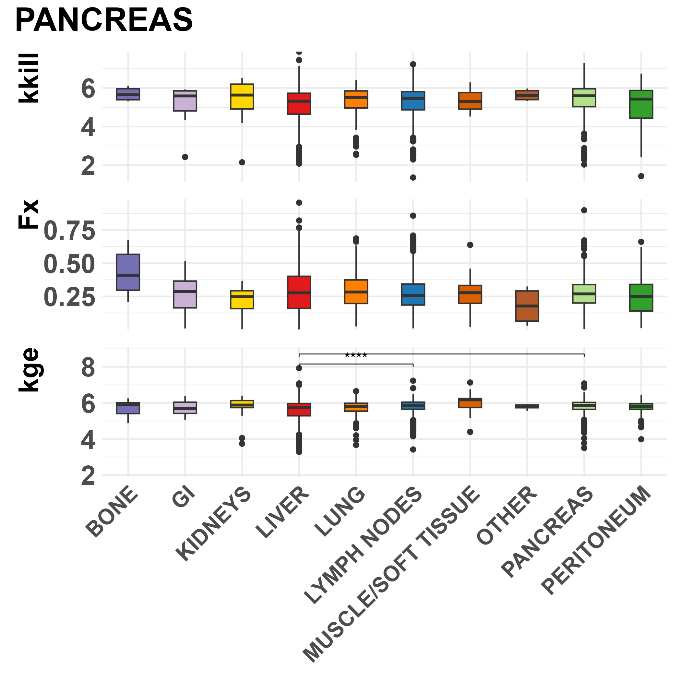

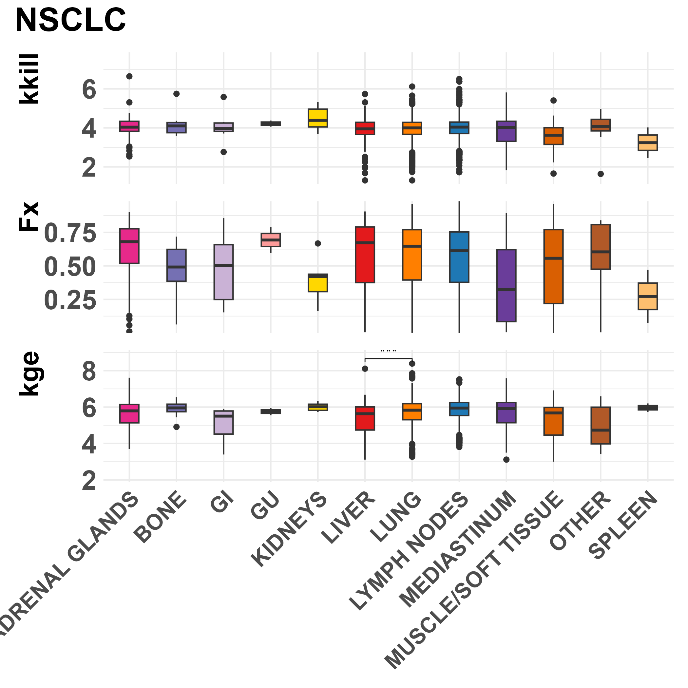

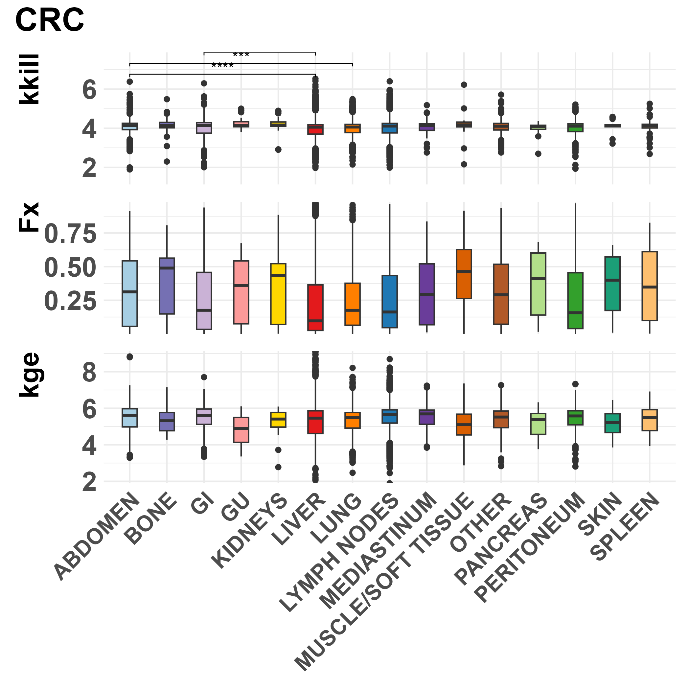

Boxplots show distinct differences for lesion growth parameters across organs within metastatic cancer cohorts. Asterisks (*) indicate statistically significant differences by one-way ANOVA (p<0.05).

Supplement Figure 5

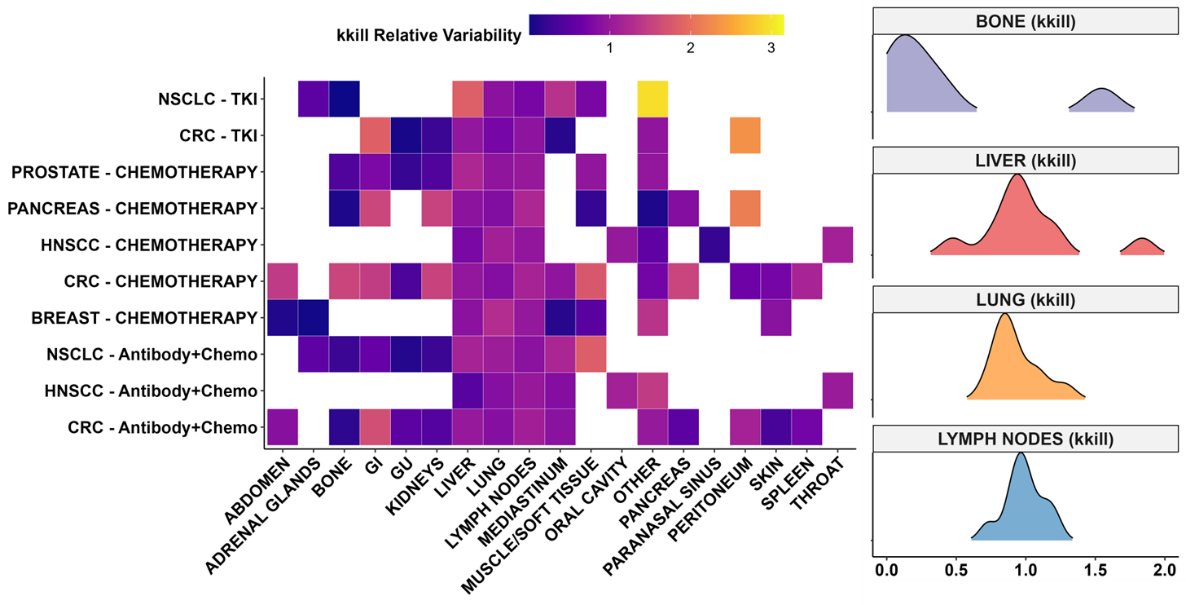

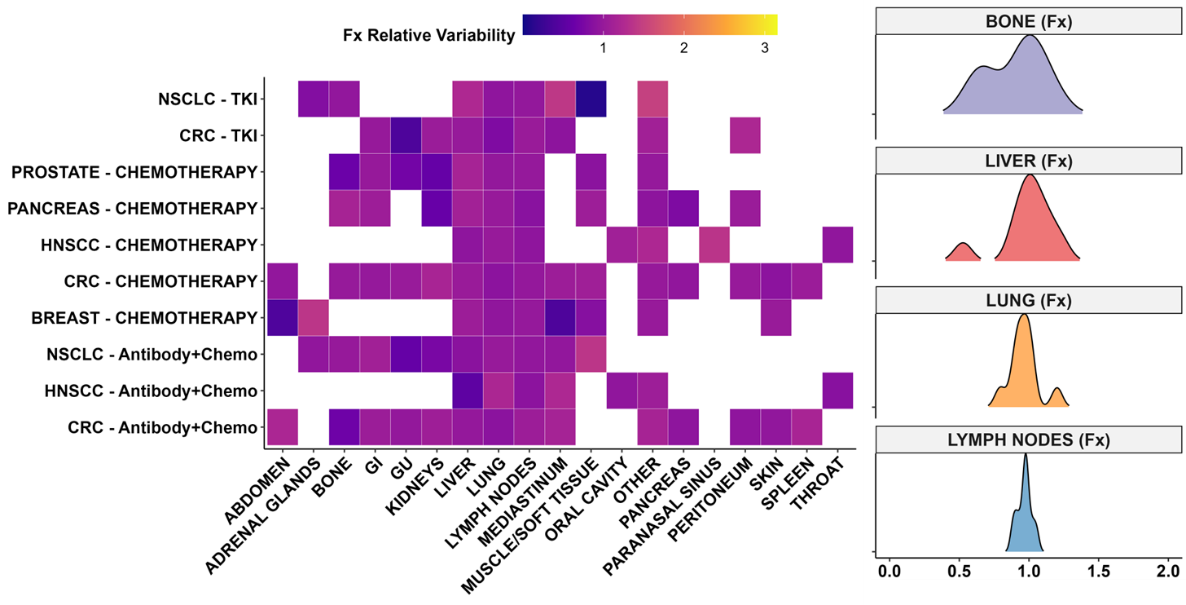

The other two TGI parameter heatmaps demonstrate the variability of across lesion locations within and across the different cancer:treatment groups. The resistant fraction (Fx) saw the least variability compared to the kkill and kge parameters.

Supplement Figure 6

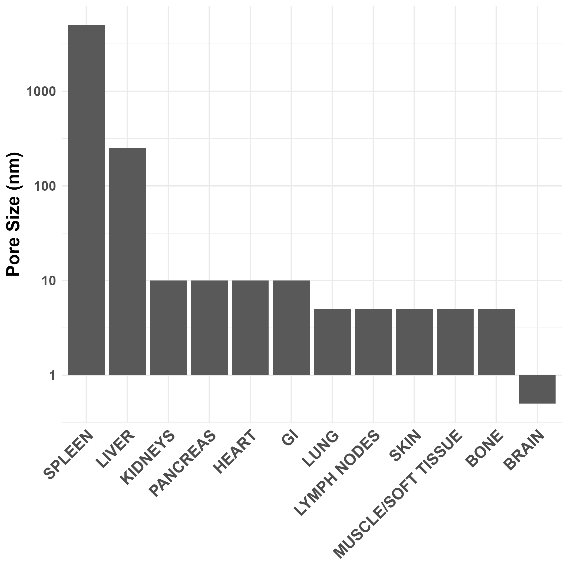

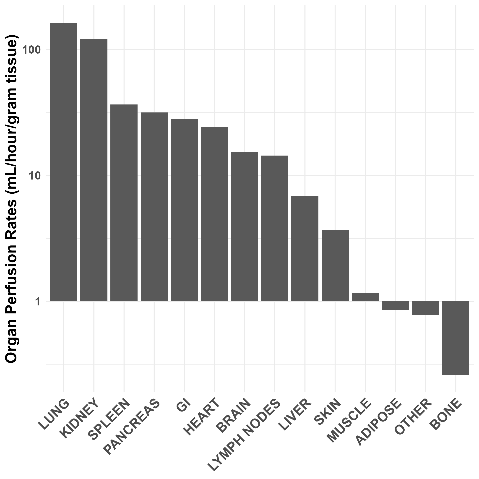

Top chart displays capillary fenestration size across key organ sites and bottom chart displays organ perfusion data (mL/hr/gm tissue). These attributes were factored together to create the VaPLI score for each organ/location described in the Methods.
